# Commercial Serology Assays Predict Neutralization Activity Against SARS-CoV-2

**DOI:** 10.1101/2020.07.10.20150946

**Authors:** Raymond T. Suhandynata, Melissa A. Hoffman, Deli Huang, Jenny T. Tran, Michael J. Kelner, Sharon L. Reed, Ronald W. McLawhon, James E. Voss, David Nemazee, Robert L. Fitzgerald

## Abstract

**Background:** Currently it is unknown whether a positive serology results correlates with protective immunity against SARS-CoV-2. There are also concerns regarding the low positive predictive value of SARS-CoV-2 serology tests, especially when testing populations with low disease prevalence.

**Methods:** A neutralization assay was validated in a set of PCR confirmed positive specimens and in a negative cohort. 9,530 specimens were screened using the Diazyme SARS-CoV-2 IgG serology assay and all positive results (N=164) were reanalyzed using the neutralization assay, the Roche total immunoglobin assay, and the Abbott IgG assay. The relationship between the magnitude of a positive SARS-CoV-2 serology result and the levels of neutralizing antibodies detected was correlated. Neutralizing antibody titers (ID50) were also longitudinally monitored in SARS-CoV-2 PCR confirmed patients.

**Results:** The SARS-CoV-2 neutralization assay had a PPA of 96.6% with a SARS-CoV-2 PCR test and a NPA of 98.0% across 100 negative controls. ID50 neutralization titers positively correlated with all three clinical serology platforms. Longitudinal monitoring of hospitalized PCR confirmed COVID-19 patients demonstrates they made high neutralization titers against SARS-CoV-2. PPA between the Diazyme IgG assay alone and the neutralization assay was 50.6%, while combining the Diazyme IgG assay with either the Roche or Abbott platforms increased the PPA to 79.2% and 78.4%, respectively.

**Conclusions:** For the first time, we demonstrate that three widely available clinical serology assays positively correlate with SARS-CoV-2 neutralization activity observed in COVID-19 patients. When a two-platform screen and confirm approach was used for SARS-CoV-2 serology, nearly 80% of two-platform positive specimens had neutralization titers (ID50 >50).

**Summary:** Clinical performance of a SARS-CoV-2 neutralization assay was evaluated using SARS-CoV-2 PCR confirmed patients and SARS-CoV-2 negative individuals. The neutralization assay was compared with results from SARS-CoV-2 positive serology specimens. We demonstrate that positive SARS-CoV-2 serology results correlate with the presence of neutralization activity against SARS-CoV-2. We show a high false positive rate when using a single SARS-CoV-2 serology platform to screen populations with low disease prevalence; and confirm that using a two-platform approach for COVID-19 seropositives greatly improves positive predictive value for neutralization.

## Introduction

The 2019 coronavirus pandemic (COVID-19) is caused by the highly pathogenic severe acute respiratory syndrome-related corona virus 2 (SARS-CoV-2) that was first discovered in Wuhan, China (1,2). As governments across the world struggle to mitigate the spread of the virus, the world-wide economy has been greatly impacted by the resulting shutdown and social distancing protocols that have been implemented (3–7). As nations begin to “re-open their economies” in stages, a major question that surrounds COVID-19 antibody testing is whether they can be used to identify individuals with protective immunity against the virus (8). Neutralizing antibodies (Nabs) play a key role in the quest for protective immunity against SARS-CoV-2 (9), and ID50 neutralization titers have been the gold standard to assess protective immunity against smallpox, polio, and influenza viruses following vaccine administration (10–12). A number of reports have characterized the clinical performance of commercially available SARS-CoV-2 serology assays (13–19). However, whether commercial SARS-CoV-2 serology platforms correlate with the presence of neutralizing antibodies and protective immunity against COVID-19 still needs further exploration (20–26).

Jääskeläinen et al., recently reported a comparison of six SARS-CoV-2 immunoassays and a microneutralization assay against SARS-CoV-2 (27) and demonstrated that 41 of 62 COVID-19 patients showed neutralizing antibodies. Our study evaluates the sensitivity and specificity of a pseudovirus based neutralization assay and investigates the correlation between the neutralization assay (28,29) and SARS-CoV-2 serology platforms in a large cohort of clinical specimens. For the first time, we determined the orthogonality of the Diazyme, Roche, and Abbott assays to initiate a dual immunoassay approach for confirming positive SARS-CoV-2 serology results, as suggested by the Centers for Disease Control and Prevention (CDC) (30). This screen and confirm approach was applied retrospectively to 9,530 SARS-CoV-2 tests and for the first time we demonstrate that combining two orthogonal serology assays significantly improves the predictive value for identifying neutralizing antibodies.

## Materials and Methods

### Study Design and Patient Cohort

The patient cohorts used in this study are illustrated in **Figure 1**. Briefly, the main cohort (**Figure 1A**) consisted of 9,530 consecutive specimens (K-EDTA, lithium-heparin plasma separator tubes, and/or serum separator tubes) that were screened at UC San Diego Health using the Diazyme IgG assay from 4/14/20 until 6/12/20. The specimens that were positive using the Diazyme IgG assay (N=164) were separated into subgroups depending on symptoms, if they were admitted, or if they were from a skilled nursing facility (SNF). All of the specimens that screened positive with the Diazyme assay were retrospectively tested using the neutralization, Roche, and Abbott assays. A separate cohort (**Figure 1B**) was used to evaluate the relationship between hospitalized SARS-CoV-2 PCR confirmed positive patients (31 patients, 87 samples) and the neutralization assay. The last cohort (**Figure 1C**) was used to evaluate the relationship between SARS-CoV-2 negative samples (100 samples) and the neutralization assay. The negative cohort consisted of 7 patients positive for other coronaviruses (229E, HKU1, NL63, or OC42), 4 Rhinovirus positive patients, 10 apparently healthy individuals, and 79 COVID-19 naïve samples (collected in 2018, stored −20 ^°^C).

**Figure 1:**
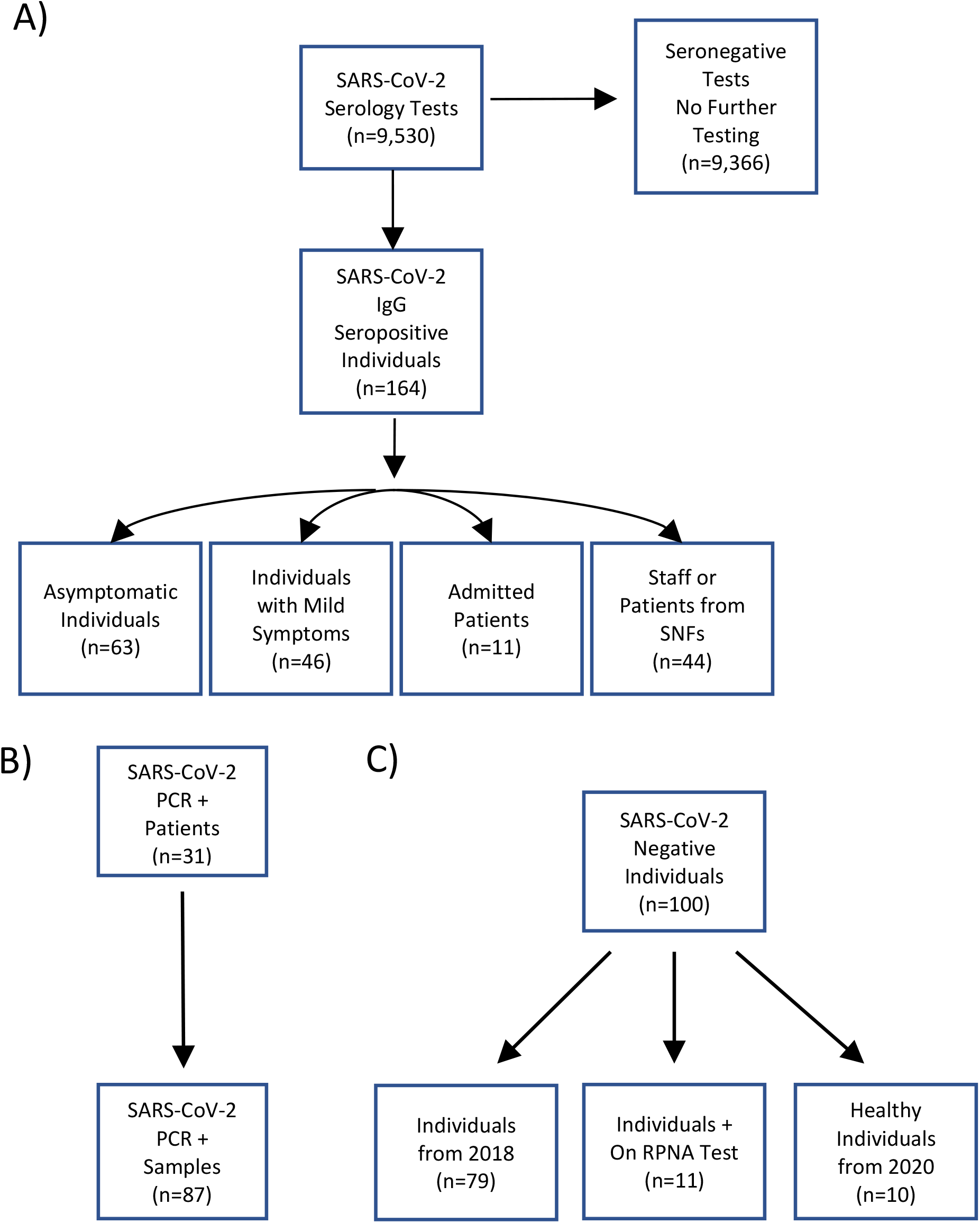
Summary of Cohorts Used. Flowcharts of the cohorts used for **A)** Main Cohort, **B)** SARS-CoV-2 PCR positive cohort, and **C)** SARS-CoV-2 negative cohort.

**Figure 2:**
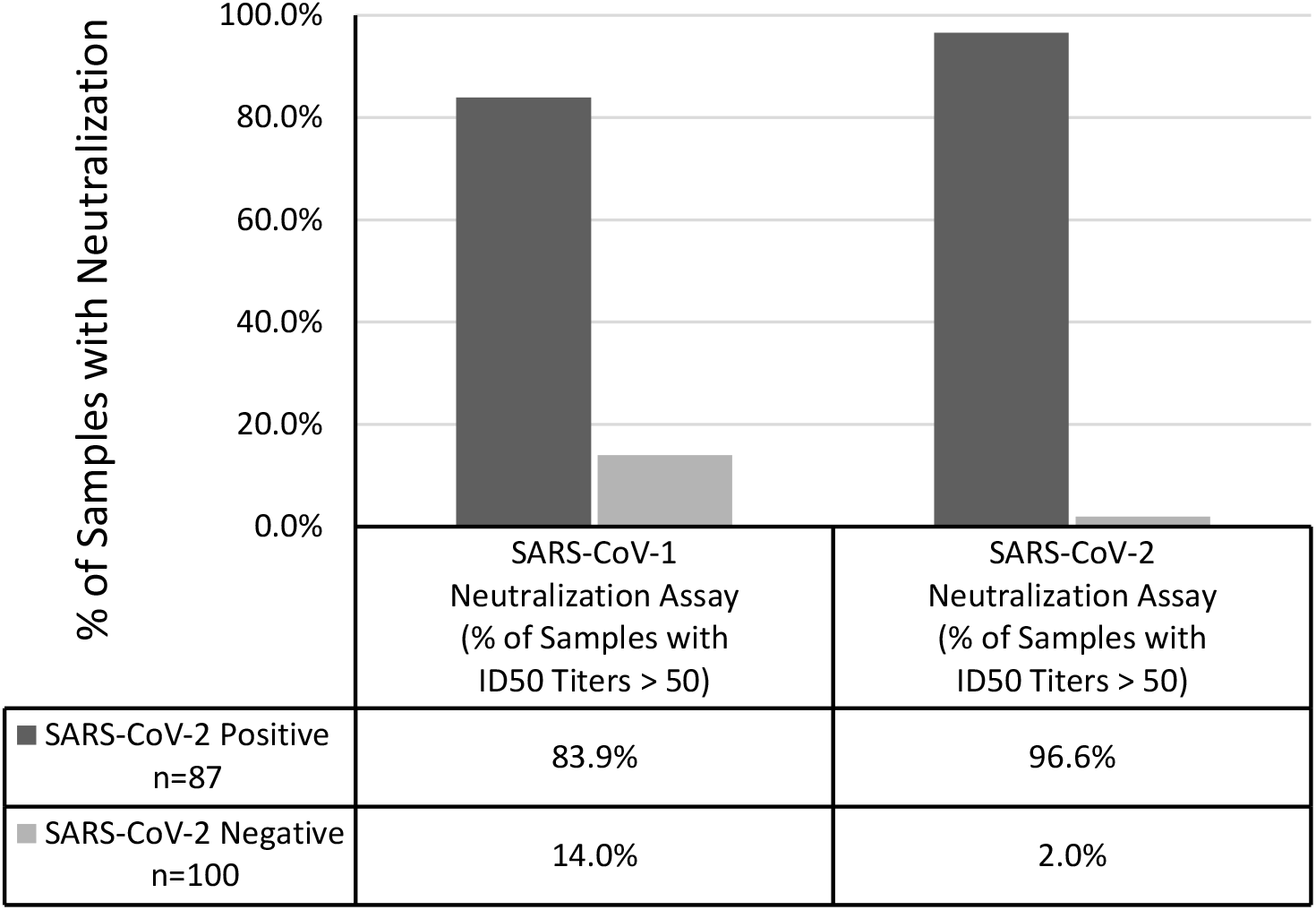
COVID-19 Patients Produce Neutralization Activity Against SARS-CoV-2 and SARS-CoV-1. Percentage of samples from SARS-CoV-2 PCR confirmed patients (dark grey) and a SARS-CoV-2 negative patient cohort (light grey) that had ID50 titers >50 against SARS-CoV-2 or SARS-CoV-1.

**Figure 3:**
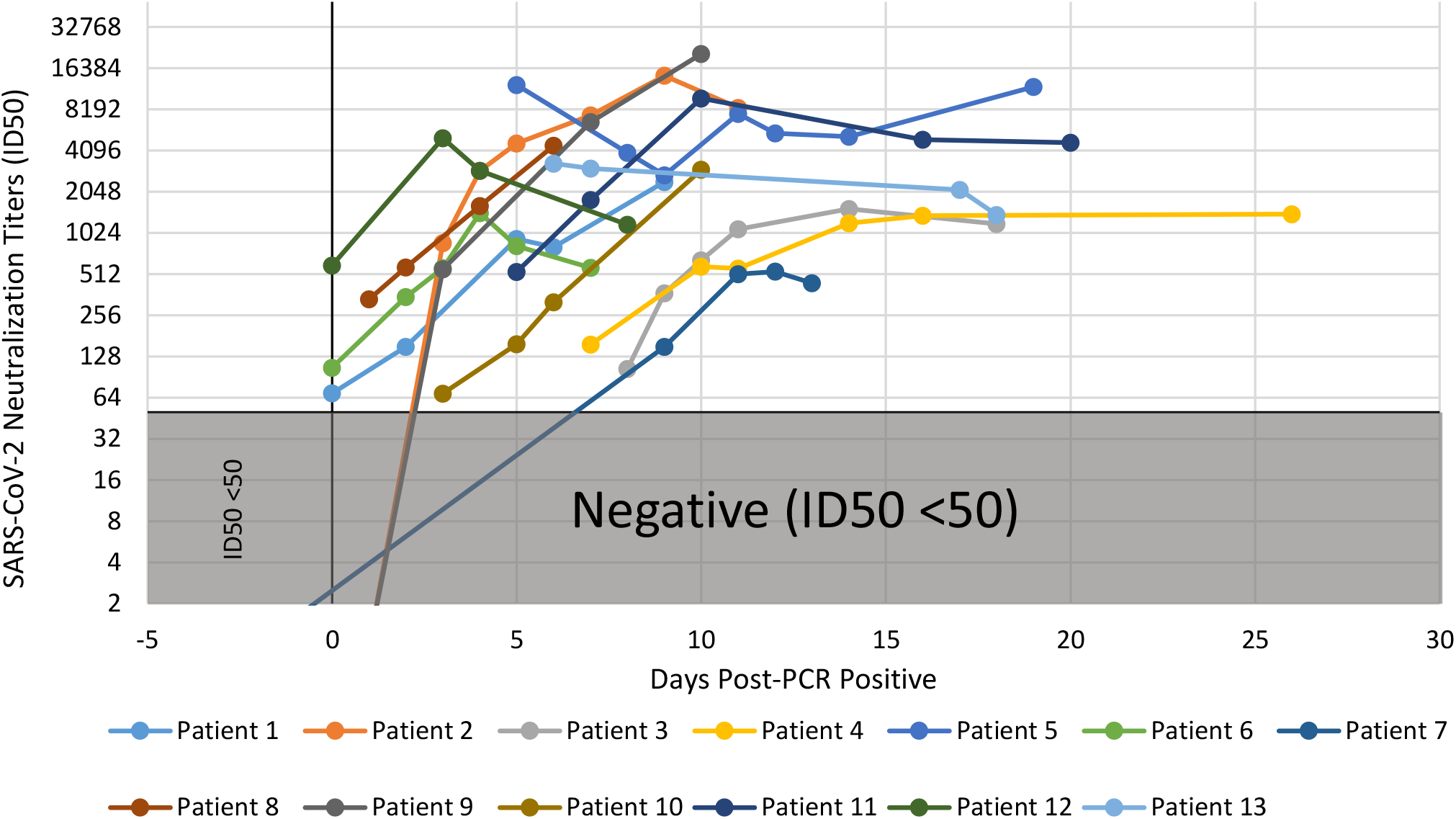
Longitudinal Monitoring of SARS-CoV-2 Neutralization Titers in COVID-19 Patients. The ID50 titers of 13 SARS-CoV-2 PCR confirmed patients are plotted on a semi-log scale (Y-axis) with the number of days following a positive PCR result indicated for each sample (X-axis). ID50 values of <50 are considered negative for neutralization activity and represented by the greyed-out area.

**Figure 4:**
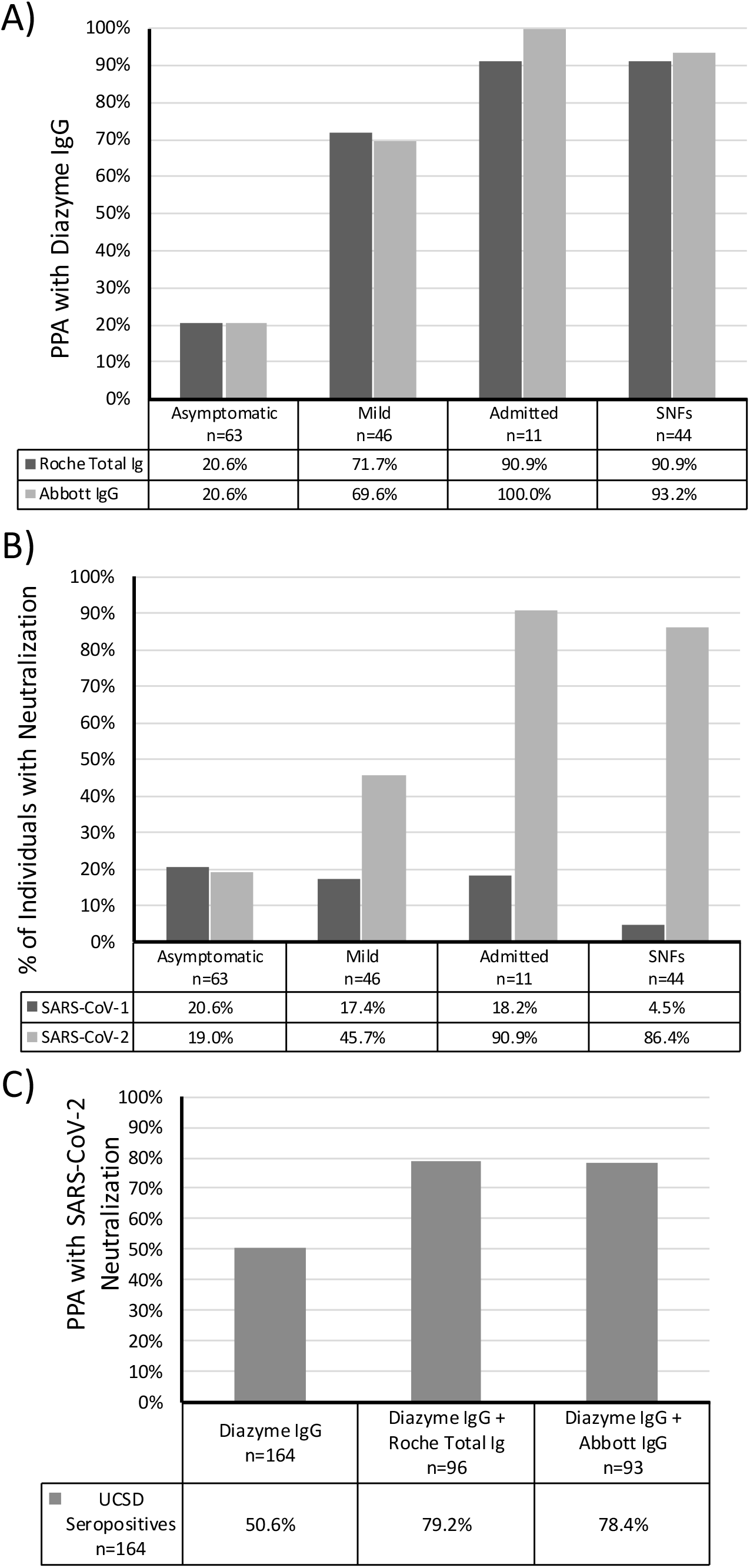
Interplatform agreement between serology and neutralization activity. The PPA of between the Diazyme IgG and Roche or Abbott assays are shown for 4 different populations of individuals. **B)** The percentage of individuals with neutralization activity against SARS-CoV-2 or SARS-CoV-1 are shown for 4 different populations of individuals. **C)** The overall PPA between the Diazyme IgG assay alone and in combination with the Roche or Abbott serology platforms with SARS-CoV-2 neutralization activity are indicated for 164 seropositive individuals identified on the Diazyme IgG platform. The number individuals in each group are indicated in the corresponding tables in **Figures A-C**.

The group (N=251) used to evaluate a cutoff for neutralization activity on each of the commercial serology platforms, shown in **Figure 5**, included the 164 samples that were seropositive for IgG on the Diazyme platform and the 87 specimens from the SARS-CoV-2 PCR confirmed patients (who were also positive on the Diazyme IgG assay). Samples were split into two groups (Positive and Negative) based on the presence or absence of SARS-CoV-2 neutralization activity.

**Figure 5:**
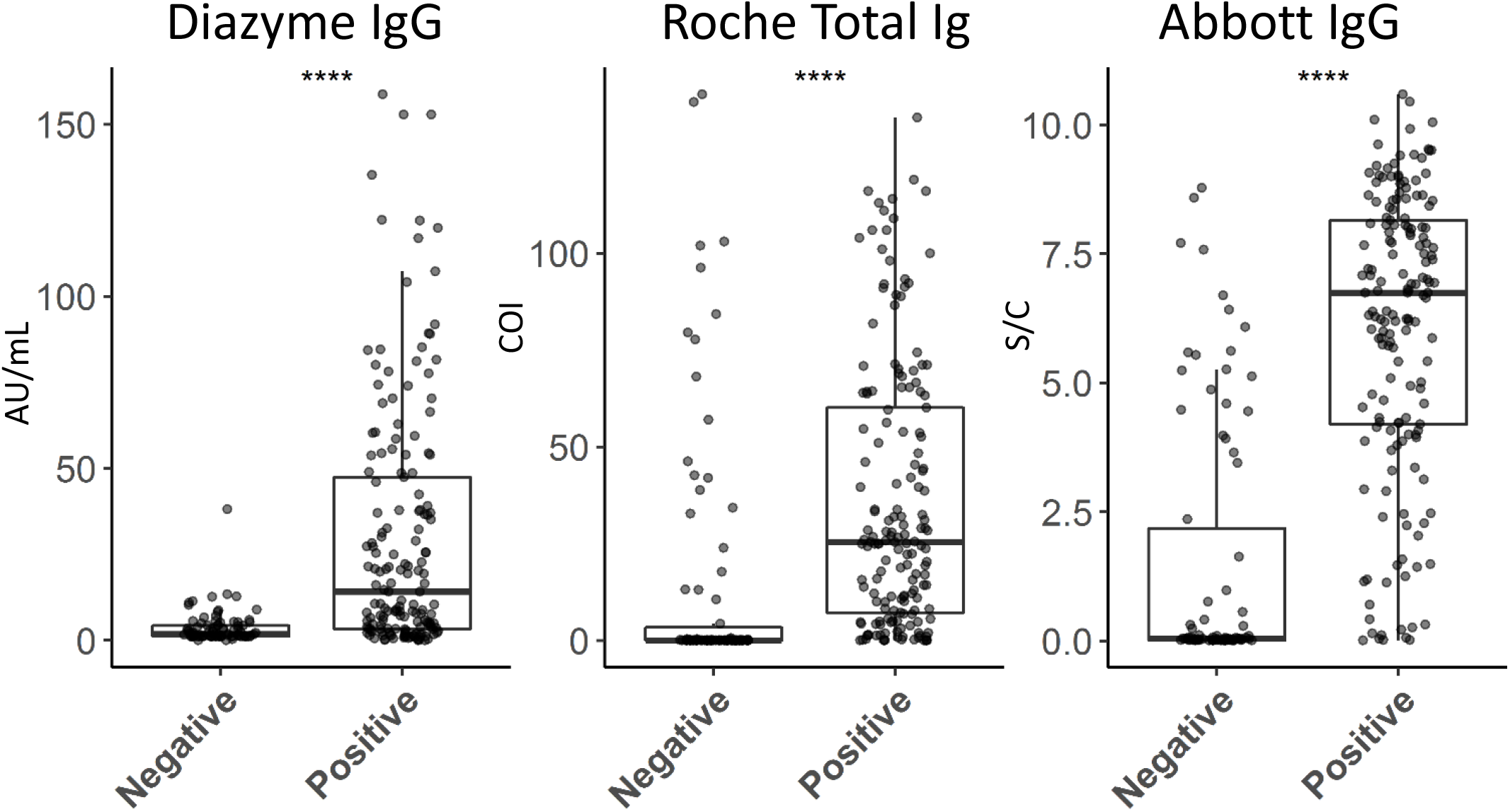
Distribution of commercial serology platform results in samples with and without SARS-CoV-2 neutralization activity that initially screened positive by the Diazyme IgG assay. AU/mL, COI, and S/C values (Y-axis) on the Diazyme IgG, Roche total Ig, and Abbott IgG platforms, respectively, are shown in boxplots (whiskers are up to but no greater than 1.5 times the IQR) for samples with (positive) or without (negative) ID50 neutralization titers against SARS-CoV-2 (X-axis). **** indicates significant difference between positive and negative group at a p value of <0.0001 by t-test.

All patient samples were collected under UCSD IRB protocol 181656.

### Confirmation of SARS-CoV-2 Positive Patients

All 31 SARS-CoV-2 patients were positively confirmed for COVID-19 by an emergency use authorized (EUA) nucleic acid test that had also been validated in our laboratory. For brevity these will be referred to as SARS-CoV-2 PCR confirmed patients.

### Serology

Serology was performed on the Diazyme DZ-Lite 3000 plus clinical analyzer as previously described for IgG (13). Serology was performed on the Roche Cobas 8000 e801 analyzer (Roche Elecsys Anti-SARS-CoV-2 total Ig) and the Abbott ARCHITECT i1000SR analyzer (Abbott SARS-CoV-2 IgG) according to the manufacturer’s instructions. Plasma (Li-Heparin or K-EDTA) and serum samples were analyzed in a manner consistent with the package inserts. The Diazyme platform reports results as absorbance units per mL (AU/mL); values ≥ 1.00 AU/mL are considered reactive. The Roche platform reports results in the form of a cutoff index (COI; signal of sample/cutoff); values ≥ 1.00 COI are considered reactive. The Abbott platform reports results in the form of an Index value (S/C); Index values ≥ 1.4 S/C are considered positive. For consistency, we refer to reactive and non-reactive to mean the same as positive and negative throughout the manuscript.

### SARS-CoV-2 and SARS-CoV-1 Pseudovirus Neutralization Assays

The neutralization assays were performed as previously described using a pseudovirus (PSV) (29). The PSV assay was established for both SARS-CoV-1 and SARS CoV-2 using murine leukemia virus (MLV) based PSV. The assay used single cycle infectious viral particles containing firefly luciferase. The amount of luminescence in HeLa cells that stably expressed the cell surface receptor angiotensin converting enzyme 2 were measured after viral infection. Titers of 50% inhibitory dilution (ID50) were determined. ID50 titers of greater than or equal to 50 was considered a positive neutralization result. The PSV assay was compared with a live replication-competent virus and neutralizing antibodies identified with the assay provided protection against high dose SARS-CoV-2 infection in a hamster animal model (29).

### GenMark ePlex Respiratory Pathogen Nucleic Acid Test (RPNA)

The RPNA test was performed as previously described to confirm patients with other respiratory pathogens (13).

### Statistical Analyses

Data was analyzed using R in Rstudio and linear regression analysis for all figures were performed in excel or Rstudio. Box and whisker plots were generated in Rstudio. Two sample t-test and Fishers exact test were performed in R for the box and whisker plots and demographics table, respectively.

## Results

### Cohort Demographics

Demographics of the Diazyme IgG seropositive individuals, that are divided based on symptom severity, are shown for age, body mass index (BMI), ethnicity, underlying medical conditions, PCR positivity for COVID-19, and positivity for neutralization activity against SARS-CoV-2 (**Table 1)**. Median (IQR) age was significantly different among all three symptom severity groups using the Fishers exact test (P-value <0.0001) with asymptomatic patients being younger. However, no significant difference was observed for BMI or gender. Significant differences in ethnicity was observed across all three groups (P-value 0.005), with 40% of the admitted group being Hispanic and 63% of the mildly symptomatic group being white. A significant difference was observed for underlying medical conditions (P-value 0.018), 60% of admitted patients had underlying conditions vs 32% in the asymptomatic group. Significant difference (P-value <0.0001) was observed in the percentage of individuals which were positive for COVID-19 by PCR, with 88% of admitted patients having a positive PCR test for SARS-CoV-2 vs 2% testing in the asymptomatic group. Significant difference (P-value <0.0001) was observed across all three groups for the presence of neutralization activity against SARS-CoV-2, with 98% of admitted patients having neutralization activity vs 19% in the asymptomatic group. Demographics for the 44 Diazyme IgG seropositive patients and staff from SNFs could not be retrieved as they were deidentified prior to analysis.

**Table 1.** Demographics of Seropositive Individuals

|  | **Asymptomatic**<br>**N = 63** | **Mild Symptoms**<br>**N = 46** | **Admitted**<br>**N = 42** | **P-value** |
| --- | --- | --- | --- | --- |
| **Age (years)** | 43 (33 - 49) | 49 (36 - 64) | 52 (43 - 68) | < 0.0001 |
| **Gender** |  |  |  | 0.21 |
| Male | 33 (52%) | 25 (54%) | 29 (69%) |  |
| Female | 30 (48%) | 21 (46%) | 13 (31%) |  |
| **BMI** |  |  |  | 0.23 |
| Median (IQR) | 25 (23 - 29) | 26 (24 - 28) | 28 (24 - 30) |  |
| Not Available | 15 (24%) | 1 (2%) | 0 (0%) |  |
| **Ethnicity** |  |  |  | 0.005 |
| Asian | 9 (14%) | 1 (2%) | 4 (10%) |  |
| Black | 5 (8%) | 0 (0%) | 3 (7%) |  |
| Filipino | 7 (11%) | 7 (15%) | 2 (5%) |  |
| Hispanic | 14 (22%) | 8 (17%) | 17 (40%) |  |
| Mixed | 3 (5%) | 0 (0%) | 1 (2%) |  |
| White | 20 (32%) | 29 (63%) | 15 (36%) |  |
| Not Available | 5 (8%) | 1 (2%) | 0 (0%) |  |
| **Underlying Medical Conditions** |  |  |  | 0.018 |
| Yes | 20 (32%) | 19 (41%) | 25 (60%) |  |
| **COVID-19 PCR** |  |  |  | < 0.0001 |
| Negative | 54 (86%) | 17 (37%) | 5 (12%) |  |
| Positive | 1 (2%) | 15 (33%) | 37 (88%) |  |
| Not Tested | 8 (13%) | 14 (30%) | 0 (0%) |  |
| **Neutralization Activity (SARS-CoV-2)** |  |  |  | < 0.0001 |
| Negative | 51 (81%) | 25 (54%) | 1 (2%) |  |
| Positive | 12 (19%) | 21 (46%) | 41 (98%) |  |

### Clinical Evaluation of a SARS-CoV-2 Neutralization Assay

The SARS-CoV-2 neutralization assay detected neutralization activity in 96.6% of SARS-CoV-2 PCR confirmed samples, and in 2.0% of the SARS-CoV-2 negative samples (**Figure 2**). 83.9% of the samples from SARS-CoV-2 PCR confirmed patients and 14.0% of the SARS-CoV-2 negative samples had SARS-CoV-1 neutralization activity (**Figure 2**). The median number of days post PCR positivity for samples from SARS-CoV-2 patients was 9 days, with an interquartile range of 5 – 15 days. SARS-CoV-1 or SARS-CoV-2 neutralization was not detected in 7 patients infected with non-COVID-19 coronaviruses and 4 patients infected with rhinovirus (**Supplementary Table 1**).

### Correlation of ID50 Neutralization Titers to Commercial SARS-CoV-2 Serology Assays

Linear regression analysis was performed to evaluate the relationship between neutralization titers (ID50) and three commercially available SARS-CoV-2 serology platforms for PCR confirmed patients (**Supplementary Figure 1)**. All three platforms showed positive correlation to SARS-CoV-2 ID50 titers.

### Longitudinal Monitoring of ID50 Neutralization Titers in COVID-19 patients

SARS-CoV-2 ID50 neutralization titers were longitudinally monitored in 13 PCR confirmed SARS-CoV-2 patients. Serial sampling revealed that SARS-CoV-2 ID50 neutralization titers increased with disease progression in 11 of the 13 patients, while the remaining two patients maintained ID50 neutralization titers of ≥ 1000 (**Figure 3**). Once elevated, neutralization titers appeared to reach a plateau that stayed elevated up to 25 days (longest time tested). Notably, all patients developed high titers by two weeks after scoring PCR positive.

### Retrospective Analysis of Diazyme Screened Seropositive Patients

The 164 Diazyme screened IgG seropositive patient samples were also analyzed on the Roche Total Ig and Abbott IgG SARS-CoV-2 serology platforms. The overall PPAs between the Diazyme IgG assay and the Roche total Ig or Abbott IgG assays were 58.5% and 59.1%, respectively (**Supplementary Figure 2**). The PPA between Diazyme and Roche was 20.6% for asymptomatic individuals, 71.7% for patients with mild symptoms, 90.9% for admitted patients, and 90.9% for patients/staff from SNFs (**Figure 4A**). The PPAs between Diazyme and Abbott were similar to what was observed with the Roche assay (**Figure 4A**). The percentage of individuals with detectable levels of SARS-CoV-2 neutralization activity increased with increasing PPA between serology platforms (**Figure 4B)**, while the percentage of individuals with detectable levels of SARS-CoV-1 neutralization activity decreased with increasing PPA between serology platforms (**Figure 4B)**. Overall, 50.6% of seropositive individuals (n=164) had detectable neutralization activity, which increased to 79.2% and 78.4% when using the Roche and Abbott platforms to confirm Diazyme seropositives, respectively (**Figure 4C**). No significant differences were observed for ID50 neutralization titers across the different groups (**Supplementary Table 2)**. All samples from the PCR confirmed positive patients with a positive serology result by either the Diazyme IgG (n=71), Roche total Ig (n=76), and Abbott IgG (n=75) assays had neutralization activity (> ID50).

### Multi-platform Cutoff Evaluation for SARS-CoV-2 Neutralization Activity

Boxplots of the observed responses on the Diazyme, Roche, and Abbott platforms, from Diazyme IgG seropositives, which were split based on the presence or absence of SARS-CoV-2 neutralization activity, are shown in **Figure 5**. The medians (IQR) for the Diazyme IgG assay were 1.9 (1.2 – 4.3) AU/mL and 14.1 (3.3 – 47.4) AU/mL for the negative and positive groups, respectively. The medians for the Roche total Ig assay were 0.1 (0.09 – 3.4) COI and 25.5 (7.5 – 63.6) COI for the negative and positive groups, respectively. The medians for the Abbott IgG assay was 0.04 (0.02 – 1.1) S/C and 6.8 (4.2 – 8.2) S/C for the negative and positive groups, respectively. The median positive group value observed for each assay was explored as a predictive cutoff for neutralization activity. When the median value serology response was used, 98.8%, 95.1%, and 83.5% of the Diazyme, Abbott, and Roche seropositive samples above the cutoff value had detectable levels of SARS-CoV-2 neutralization activity (> ID50).

## Discussion

The SARS-CoV-2 neutralization assay had a PPA of 96.6% with a SARS-CoV-2 PCR test (n=87) and an NPA of 98.0% across 100 negative controls. Interestingly, 83.9% of samples from SARS-CoV-2 PCR confirmed patients had neutralizing activity for SARS-CoV-1 that was responsible for the original 2002-2004 SARS epidemic (31–34). This supports the finding that SARS-CoV-2 infected individuals produce neutralizing antibodies that cross-react with SARS-CoV-1 (35). Likewise, neutralization activity against SARS-CoV-1 was observed in 14.0% of COVID-19 negative individuals and could be a result of past exposure to SARS-CoV-1, lower specificity of the SARS-CoV-1 neutralization assay, or greater cross-reactivity to other common coronaviruses. The high agreement between a positive SARS-CoV-2 PCR result and the SARS-CoV-2 neutralization assay suggests that neutralization antibodies are made quickly in response to viral infection. Overall, the SARS-CoV-2 neutralization assay had PPA and NPA similar to published results for clinically validated SARS-CoV-2 serology assays [13] and provides a direct approach to assess protective immunity against SARS-CoV-2.

Regression analysis of SARS-CoV-2 neutralization titers in SARS-CoV-2 PCR confirmed positive patients correlated with the corresponding values on the Diazyme, Roche, and Abbott platforms. The Diazyme IgG assay had the strongest correlation with SARS-CoV-2 neutralization titers, and could be a result of the linear characteristics of this platform (13). Furthermore, **Supplementary Figure 1** illustrates that all platforms were highly sensitive for the detection of ID50 neutralization titers greater than 1000. 98.9% of Diazyme IgG, and 100% for both Roche and Abbott platforms were seropositive when the ID50 titers were above 1000. Longitudinal monitoring of SARS-CoV-2 ID50 neutralization titers in PCR confirmed SARS-CoV-2 PCR positive hospitalized patients revealed that all patients developed robust neutralization titers during the course of infection (**Figure 3**). These results suggest that these commercial serology tests correlate with neutralization and imply protective immunity against SARS-CoV-2. These results open avenues for eventually monitoring patient immune response against vaccines that are being developed for COVID-19.

Using the Roche or Abbott SARS-CoV-2 serology platforms to confirm specimens initially screened positive on the Diazyme IgG assay demonstrated that nearly 40% of the 164 IgG seropositive individuals would have been classified as false positives by the confirmatory assay. As predicted previously (13), the percentage of individuals that were retrospectively identified as falsely positive depended greatly on the population of individuals being tested. The retrospective false positive rate was as high as 79.4% in asymptomatic individuals, approximately 30% in individuals with mild symptoms, and potentially 0% in admitted patients. This underlines the effect of disease prevalence, and the dangers of screening low prevalence populations even when using a highly specific SARS-CoV-2 serology assay.

100% of specimens that tested positive on all three commercially available serology platforms had detectable ID50 titers (> 50) against SARS-CoV-2. In contrast, only 50.6% of the 164 seropositive individuals identified on the Diazyme platform had neutralization titers (**Figure 4C)**, a percentage similar to the PPA between Diazyme and the Roche and Abbott platforms (**Figure 4A**). However, using the Roche or Abbott platforms to confirm IgG seropositive individuals, would have resulted in nearly 80% of reported seropositive individuals also having neutralization activity against SARS-CoV-2 (**Figure 4C**).

Interestingly, no difference was observed in the SARS-CoV-2 ID50 titers across the four different populations that were tested (**Supplementary Table 2**), suggesting that individuals make comparable levels of neutralization antibodies regardless of symptoms. The finding that 80% of two-platform confirmed positive serology results have neutralizing activity is an important finding, addressing a major concern when using COVID-19 serology to screen low disease prevalence populations. However, roughly 20% of the two-platform confirmed seropositive individuals did not have SARS-CoV-2 neutralization activity and suggests that a clinically validated SARS-CoV-2 neutralization assay is required for understanding if patients have protective immunity against COVID-19.

In principle, not all COVID-19 patients should have neutralizing antibodies, as these take time to develop after infection, and neutralizing antibodies represent a small subset of antibodies produced [29]. It is therefore encouraging that virtually all SARS-CoV-2 seropositive individuals produce neutralizing antibodies. The presence of SARS-CoV-2 neutralizing antibodies patients with symptoms of cytokine storm may encourage more aggressive steroid treatment in those patients.

We explored using the median positive serology result as the cutoff value for determining the presence of neutralizing antibodies (**Figure 5**). Interestingly, specimens with serology results above the median were highly associated with neutralizing antibodies. 98.9, 95.1 and 83.5% of samples above the median serology value contained neutralizing antibodies with ID50 titers greater than 50 when using the Diazyme, Abbott and Roche assays respectively.

We observed significant differences in the demographics of 151 Diazyme seropositive individuals that were asymptomatic, mildly symptomatic, and hospital admitted. In particular, although the percentage of Hispanic individuals comprised 25.8% of this population, they accounted for 40% of the hospital admitted cases; of which 98% were PCR positive for COVID-19. This is similar to another recent report which observed that 60% of all pregnant women hospitalized for COVID-19 were of Hispanic ethnicity (36).

The retrospective analysis of only the 164 seropositive SARS-CoV-2 individuals and not the population of 9530 individuals is a limitation of the study, preventing a fair comparison of the Diazyme platform with the Roche and Abbott SARS-CoV-2 serology platforms. This is because these individuals were effectively pre-screened for SARS-CoV-2 seropositivity before analysis on either the Roche or Abbott platforms. Moreover, out of the 9,530 tests performed, the estimated overall false positive rate of the Diazyme IgG assay was between 0.7 - 0.9%, and is equal to or lower than its published value of 0.9% (13). This suggests that our findings would have been similar if either the Roche total Ig or Abbott IgG assays were used as the screening platform and not the confirmatory platform, as both assays have been reported to have specificity that exceeds 99% (14–16); underlining the dangers of using any one single serology platform to screen low prevalence populations for SARS-CoV-2 serology.

We report a comprehensive retrospective study of 164 SARS-CoV-2 seropositive samples from 9,530 SARS-CoV-2 serology tests and for the first time demonstrate that serology results correlate with neutralization. Our study demonstrates the risk of using a single serology platform to identify SARS-CoV-2 seropositive individuals in low prevalence populations and highlights the benefits of a two-platform approach. Finally, the inclusion of serology and neutralization activity found in mild and asymptomatic cases of COVID-19 are also unique, as cohorts used for other reports are generally of hospitalized patients.

## Data Availability

All data is stored on UCSD password protected servers.

## Acknowledgments

We thank all of the staff in the UC San Diego Health clinical laboratories for their help identifying specimens for sensitivity and specificity testing. We also thank Amy Rockefeller and Ernestine Ferrer for valuable technical expertise. We would like to thank Waters Corporation (RTS) and Roche Diagnostics (MAH) for supporting the clinical chemistry fellowship at the University of California-San Diego.

